# A Population-based Study of Sex Differences in Cognitive Impairment and Dementia Among Incident Atrial Fibrillation Patients and the Influence of Thromboprophylaxis

**DOI:** 10.64898/2026.07.22.26358739

**Authors:** Karan Saraf, Anamaria Savu, Lena Rivard, Padma Kaul, Roopinder K Sandhu

**Affiliations:** Libin Cardiovascular Institute, University of Calgary, Calgary, AB, Canada; Division of Cardiology, University of Alberta, Edmonton, AB, Canada; The Canadian VIGOUR Center, Edmonton, AB, Canada; Montreal Heart Institute, Montreal, QC, Canada

## Abstract

**Background:** It is unclear if sex differences exist in cognitive impairment and dementia associated with atrial fibrillation (AF) and whether this relationship is influenced by oral anticoagulation (OAC) therapy.

**Methods:** We identified patients ≥40 years with incident non-valvular AF (NVAF) and without previous history of cognitive impairment or dementia, stroke/transient ischemic attack or contraindication to OAC between April 1st, 2012 and March 31st, 2024, using linked administrative databases in Alberta, Canada. Outcomes included cognitive impairment, Alzheimer’s disease, vascular dementia, and all-cause mortality. Survival models were used to estimate association between sex and outcomes, and between OAC therapy and outcome stratified by sex.

**Results:** Of 66,885 patients, 43% were female. Compared to males, females were older (74 vs 68 years), frailer (14.7% vs 10.1%), and more likely to have CHA_2_DS-VA score ≥2 (73% vs 65%). OAC was initiated in 60% of both sexes within 120 days of diagnosis. Females had a higher risk for cognitive impairment (aHR 1.14 [95% CI 1.07-1.20], p<0.0001), but a lower risk of vascular dementia (aHR 0.75 [95% CI 0.63-0.90], p=0.0016) and all-cause mortality (aHR 0.91 [95% CI 0.88-0.94], p<0.001), with no difference in risk of Alzheimer’s disease (aHR 1.01 [95% CI 0.88-1.17], p=0.8496) between the sexes. Non-vitamin-K OAC (NOAC) use was associated with a lower risk of cognitive impairment compared to either warfarin or no OAC in both sexes, but a lower risk of Alzheimer’s and vascular dementia in males only.

**Conclusion:** In this large population-based study, we found females have a higher risk of cognitive impairment, but a lower risk of vascular dementia and all-cause mortality compared to males. For both sexes, NOAC use was associated with a lower risk of cognitive impairment compared to no OAC and a lower risk of Alzheimer’s and vascular dementia in males only.

## Introduction

Atrial fibrillation (AF), the most common clinical arrhythmia, has been shown to be associated with cognitive impairment and dementia.^1–3^ Proposed mechanisms include overt and silent ischemic stroke, and cerebral microinfarcts beyond the resolution of conventional neuroimaging, all of which are increased in the presence of AF.^2, 4, 5^ Consequently, there are suggestions from observational studies that oral anticoagulation (OAC) therapy is associated with reduced cognitive impairment.^6–9^

Females have been demonstrated to have higher risk of stroke and thromboembolic complications than males,^10–13^ Prior studies have also reported potential disparities in treatment, including lower rates of OAC use in females in some settings.^12–14^ There is a paucity of data examining the relationship between sex and cognitive decline and dementia with findings demonstrating that females may experience a greater burden of cognitive decline and more rapid progression to dementia, while data evaluating how this risk is associated with sex-differences in OAC therapy are lacking.^15, 16^ Accordingly, we sought to evaluate sex differences in cognitive impairment and dementia among patients with incident AF, and to examine the influence of OAC therapy on these outcomes in a large population-based cohort.

## Methods

### Study design and data sources

We conducted a retrospective population-based cohort study using linked administrative health databases from the province of Alberta, Canada. Alberta has a publicly funded health care system that provides universal access to physician and hospital services for all residents. Health services data are routinely captured through provincial administrative databases and can be deterministically linked using unique patient identifiers.

Data sources included physician billing claims, hospital discharge abstracts, ambulatory care (including emergency department) records, provincial health care insurance registry, pharmaceutical dispensing records, vital statistics, and census data. These datasets contain information on patient demographics, diagnoses, procedures, health care encounters, medication dispensations and material deprivation index at the neighborhood level based. Diagnoses are coded within hospital and ambulatory records using the International Classification of Diseases, Tenth Revisions (ICD-10), while physician claims are coded using ICD, Ninth Revision (ICD-9) (**Supplementary Table 1**).

### Study Cohort

The study cohort included adult residents of Alberta aged 40 years or older enrolled in the Alberta Health Care Insurance Plan with incident non-valvular atrial fibrillation (NVAF) identified between April 1st, 2012 and March 31st, 2024. The index date was defined as the date of the first qualifying AF diagnosis (discharge date of a hospitalization, discharge date of an emergency department visit, the latter of 2 physician claims at least 30 days apart within a 12-month window). The following patients were excluded from the cohort: 1) patients with prevalent AF, i.e., those with AF diagnosis codes in inpatient or outpatient records between April 1^st^, 2002 and March 31^st^, 2012; 2) patients who died during the index episode in which AF was diagnosed; 3) patients with valvular AF (defined as those with a history of valvular heart disease or who had undergone valvular interventions); 4) patients with contra-indications for OAC (cranial hemorrhage or major bleeding); and 5) patients with a diagnosis of cognitive impairment of any kind prior to the index date to ensure that outcomes represented incident cognitive impairment occurring after AF diagnosis. Patients were followed until death, departure from the province, or the end of the study period (March 31, 2024).

### Exposures

The primary exposure of interest was biological sex, categorized as female or male. A secondary exposure was initiation of OAC therapy within 120 days following AF diagnosis. OAC exposure was categorized into three mutually exclusive groups: no OAC, vitamin K antagonist (warfarin), or non-vitamin K oral anticoagulant (NOAC). NOACs included dabigatran, rivaroxaban, apixaban, and edoxaban. Medication exposure was determined using pharmacy dispensing records. Patients were classified according to the first OAC dispensed within the 120-day exposure window.

### Measurements

Comorbidities were defined based on the presence of previously validated ICD codes in hospitalization, emergency department visit, and physician office visit records (Supplementary Table 1).^17^ A comorbidity was considered to be present if it was coded at the index visit or healthcare visits in the previous five years, or if it was coded in two physician claims more than 30 days but less than one year apart in the previous five years. The Charlson Comorbidity Index and CHA_2_DS_2_-VASc Score were calculated at baseline. Hospital Frailty Risk Score was calculated based on comorbidities identified using hospitalization data in the 24 months prior to index date.^18^

### Outcomes

Primary outcomes included cognitive impairment, Alzheimer’s disease, vascular dementia. Outcomes were considered to have occurred if they were coded in at least one hospitalization or emergency department visit, or three physician claims at least 30 days apart within a 24-month window. Secondary outcome of interest was all-cause mortality.

### Statistical analyses

Continuous variables were summarized using median and IQR and compared using Kruskal-Wallis test. Categorical variables were summarized using frequencies and percentages and compared using chi-square tests. All-cause mortality was estimated using the Kaplan-Meier approach, expressed as 1 - survival to show the cumulative incidence of death over time. For neurocognitive events, the cumulative incidence function (CIF) was calculated to account for the competing risk of death.

Time-to-event analyses were performed using Fine-Gray proportional sub-distribution hazard models for neurocognitive outcomes and Cox proportional hazards regression models for death to estimate hazard ratios (HR) for associations between sex and outcomes.

Two primary models were constructed. The unadjusted model included sex (male, female) and OAC category (none, warfarin, NOAC), while the adjusted model included age (40-60, 60-75, 75+ years), CHA_2_DS_2_-VASc (<2, ≥2) Charlson Comorbidity Score (0, 1, 2+), Hospital Frailty Risk Score (<5, ≥5), rural residence (no, yes) and quintiles of a material deprivation score.

To evaluate whether the association between OAC therapy and outcomes differed by sex, additional models were constructed to include an interaction between sex and OAC therapy. In these models, adjusted hazard ratios for outcomes associated with warfarin and NOAC use were estimated relative to no OAC therapy. Confounding was addressed through multivariable adjustment for demographic, clinical, and socioeconomic variables listed above. Analysis was carried out using SAS software, Version 9.4. The study was approved by the University of Alberta research ethics board (Pro00010852). The ethics panel determined that the research is a retrospective database review for which subject consent for access to personally identifiable health information would not be reasonable, feasible, or practical.

## Results

The study cohort included 66,885 individuals with incident NVAF during the study period (**Table 1** and **Supplementary Figure 1**) with a median/mean follow-up time of 3.9/4.5 years. Of these, 28,465 (42.6%) were female and 38,420 (57.4%) were male. Compared to males, females were older (74 [65-82] vs 68 [59-77] years, p<0.0001), had higher prevalence of hypertension (65.4% vs 59.7%, p<0.0001), frailty (14.7% vs 10.4%, p<0.0001) and higher CHA₂DS₂-VASc score (4 [2-4] vs 2 [1-3], p<0.0001).

**Table 1.** Baseline Characteristics by sex.

| Characteristic | Female | Male | p-value |
| --- | --- | --- | --- |
| Total N | 28,465 (42.6) | 38,420 (57.4) |  |
| Age (years), Median (IQR) | 74.0 (65.0, 82.0) | 68.0 (59.0, 77.0) | <.0001 |
| Age groups, 40-65 | 7,040 (24.7) | 14,899 (38.8) | <.0001 |
| 65-74 | 7,696 (27.0) | 11,725 (30.5) |  |
| 75+ | 13,729 (48.2) | 11,796 (30.7) |  |
| Heart Failure | 4,993 (17.5) | 6,526 (17.0) | 0.0602 |
| Hypertension | 18,617 (65.4) | 22,938 (59.7) | <.0001 |
| Diabetes | 6,194 (21.8) | 10,367 (27.0) | <.0001 |
| Peripheral Vascular Disease | 1,009 (3.5) | 1,808 (4.7) | <.0001 |
| Coronary Artery Disease | 4,575 (16.1) | 9,844 (25.6) | <.0001 |
| CHA<sub>2</sub>DS<sub>2</sub>-VASc Score, 0 | 0 (0.0) | 6,136 (16.0) | <.0001 |
| 1 | 3,160 (11.1) | 7,303 (19.0) |  |
| 2+ | 25,305 (88.9) | 24,981 (65.0) |  |
| CHA<sub>2</sub>DS<sub>2</sub>-VASc Score, Median (IQR) | 4.0 (2.0, 4.0) | 2.0 (1.0, 3.0) | <.0001 |
| CHA<sub>2</sub>DS<sub>2</sub>-VA Score ≥ 2 | 20,824 (73.2) | 24,981 (65.0) | <.0001 |
| Initiated an OAC within 120 days of AF diagnosis | 17,149 (60.2) | 23,344 (60.8) | 0.1786 |
| Initiated a NOAC | 13,034 (45.8) | 17,879 (46.5) | 0.0557 |
| Initiated Warfarin | 4,115 (14.5) | 5,465 (14.2) | 0.3971 |
| Prior Myocardial Infarction | 1,925 (6.8) | 4,062 (10.6) | <.0001 |
| Anemia | 3,597 (12.6) | 3,637 (9.5) | <.0001 |
| Liver | 498 (1.7) | 913 (2.4) | <.0001 |
| Cancer | 3,323 (11.7) | 5,050 (13.1) | <.0001 |
| Chronic Kidney Disease | 1,799 (6.3) | 2,366 (6.2) | 0.3919 |
| Charlson Comorbidity Index, Median (IQR) | 1.0 (0.0, 3.0) | 1.0 (0.0, 3.0) | <.0001 |
| Charlson Comorbidity Index, 0 | 12,816 (45.0) | 16,484 (42.9) | <.0001 |
| 1-2 | 7,362 (25.9) | 10,073 (26.2) |  |
| 3+ | 8,287 (29.1) | 11,863 (30.9) |  |
| Frail | 4,194 (14.7) | 3,983 (10.4) | <.0001 |
| Rural Residence | 6,635 (23.3) | 9,363 (24.4) | 0.0015 |
| Material Deprivation, 1 | 4,452 (15.6) | 6,358 (16.5) | <.0001 |
| 2 | 4,275 (15.0) | 6,072 (15.8) |  |
| 3 | 5,130 (18.0) | 7,071 (18.4) |  |
| 4 | 6,213 (21.8) | 8,229 (21.4) |  |
| 5 | 6,283 (22.1) | 8,572 (22.3) |  |
| Missing | 2,112 (7.4) | 2,118 (5.5) |  |

CHA<sub>2</sub>DS<sub>2</sub>-VASc Score assigns 1 point for Female sex. CHA<sub>2</sub>DS<sub>2</sub>-VA Score is the score proposed by ESC 2024 guidelines and does not assign 1 point for Female sex. When CHA<sub>2</sub>DS<sub>2</sub>-VA $\geq 2$ , OAC is recommended.

Rates of OAC initiation within 120 days of AF diagnosis were similar between sexes (60.2% females and 60.8% males). Among patients initiating anticoagulation, the majority received NOACs (45.8% of females and 46.5% of males, p=0.0557) with warfarin initiated in 14.5% of females and 14.2% of males (p=0.3971).

Cognitive impairment occurred in 2,684 females (9.4%) and 2,265 males (5.9%), corresponding to a cumulative incidence at 3 years of 6.4% (95% CI 6.1-6.7) for females vs 3.8% (95% 3.6-4.1) for males at 3 years (p<0.0001). Alzheimer’s disease was diagnosed in 398 females (1.4%) and 364 males (0.9%) with a cumulative incidence at 3 years of 0.8% (95% CI 0.7-1.0) for females vs 0.6% (95% CI 0.5-0.7) for males (p<0.0001). Vascular dementia occurred in 234 females (0.8%) and 296 males (0.8%) with a cumulative incidence at 3 years of 0.6% (95% CI 0.5-0.7) for females and 0.5% (95% CI 0.4-0.5), p=0.4991) for males. All-cause mortality occurred in 8,506 females (29.9%) and 10,095 males (26.3%) with a cumulative incidence at 3 years of 18.8% (95% CI 18.3-19.3) among females and 16.3% (95% CI 16.0-16.7) among males (p<0.0001; **Figure 1**).

**Figure 1 –.**
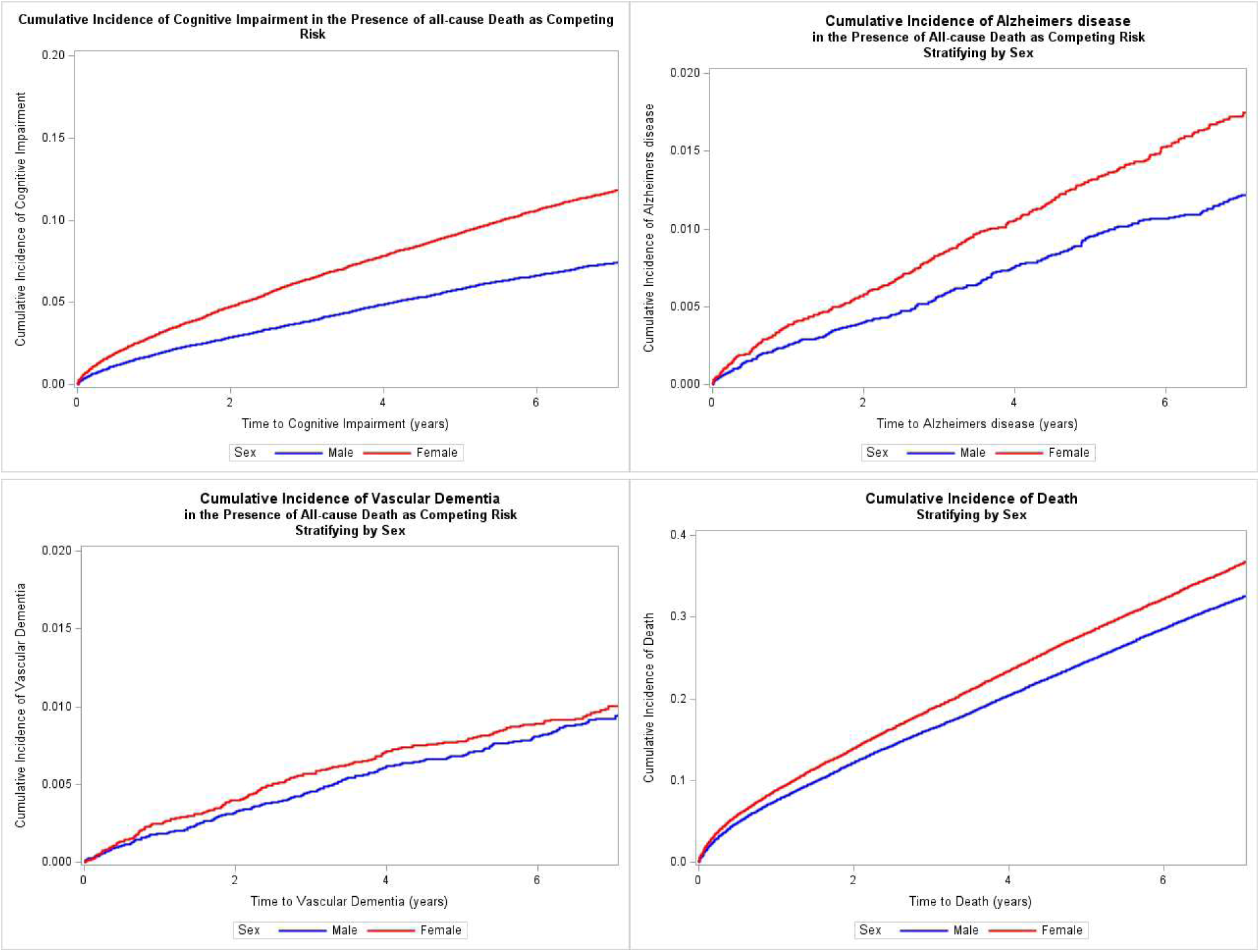
Cumulative incidence curves by sex. Compared to males, females had higher rates of cognitive impairment, Alzheimer’s disease, and all-cause mortality, while rates of vascular dementia were similar between sexes.

In unadjusted models that included sex and OAC category, females had a significantly higher risk of cognitive impairment (HR 1.63 [95% CI 1.54-1.73], p<0.0001), Alzheimer’s disease (HR 1.47 [95% CI 1.28-1.70], p<0.0001), and all-cause mortality (HR 1.16 [95% CI 1.13-1.20], p<0.0001) compared with males, while vascular dementia was similar between sexes (p=0.5020; **Table 2**). After adjustment, females retained a higher but attenuated risk for cognitive impairment (HR 1.14 [95% CI 1.07-1.20], p<0.0001), but a lower risk of vascular dementia (HR 0.75 [95% CI 0.63-0.90], p=0.0016) and all-cause mortality (HR 0.91 [95% CI 0.88-0.94], p<0.001), and no difference in risk of Alzheimer’s disease (HR 1.01 [95% CI 0.88-1.17], p=0.8496; **Table 2** and **Figure 2**).

**Figure 2 –.**
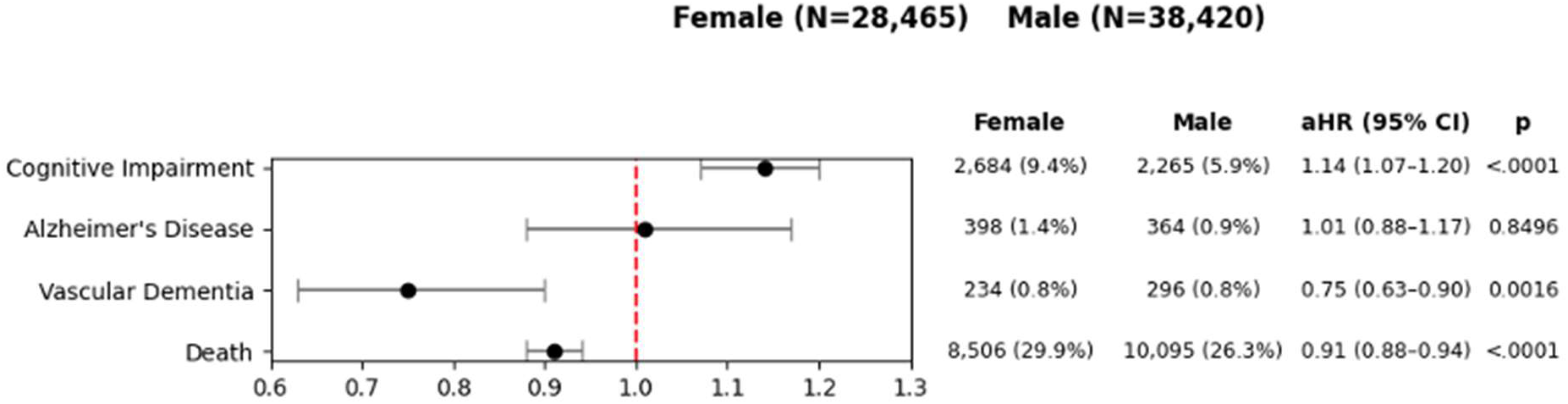
Adjusted association between sex and outcomes. After adjustment, females retained a higher but attenuated risk for cognitive impairment, but a lower risk of vascular dementia and death, with no differences in Alzheimer’s disease.

**Table 2.** Unadjusted and adjusted associations between sex and outcomes.

| Outcome | Unadjusted |  |  | Adjusted |  |  |
| --- | --- | --- | --- | --- | --- | --- |
|  | HR<br>(Female<br>vs Male) | (95% CI) | p-value | HR<br>(F vs M) | (95% CI) | p-value |
| Cognitive Impairment | 1.63 | (1.54 , 1.73) | <.0001 | 1.14 | (1.07 , 1.20) | <.0001 |
| Alzheimer's Disease | 1.47 | (1.28 , 1.70) | <.0001 | 1.01 | (0.88 , 1.17) | 0.8496 |
| Vascular Dementia | 1.06 | (0.89 , 1.26) | 0.5020 | 0.75 | (0.63 , 0.90) | 0.0016 |
| All-cause mortality | 1.16 | (1.13 , 1.20) | <.0001 | 0.91 | (0.88 , 0.94) | <.0001 |

Unadjusted models include sex and OAC (none, NOAC, Warfarin)
Adjusted models include sex, OAC (none, NOAC, Warfarin), age (40-60, 60-75, 75+), Charlson score (0, 1-2, 3+), CHA<sub>2</sub>DS<sub>2</sub>-VA $\geq 2$ (no/yes), HFRS-Frailty (no/yes), rural residence (no/yes), quintiles of material deprivation (q1-q5, missing).

In stratified analyses (**Figure 3**), among females who did not initiate OAC therapy, 1,954 of 22,632 (17.3%) developed cognitive impairment during follow-up, compared with 647 of 4,115 (15.7%) initiated on warfarin and 1,060 of 13,034 (8.1%) on NOACs. After adjustment for covariates, warfarin was associated with a higher risk of cognitive impairment compared with no OAC therapy (adjusted HR [aHR] 1.16 [95% CI 1.05-1.28]), while NOACs maintained a lower risk (aHR 0.92 [95% CI 0.84-1.00]). Among males, 1,696 of 30,152 (11.3%) without OAC therapy developed cognitive impairment, compared to 548 of 5,465 (10.0%) on warfarin, and 868 of 17,879 (4.9%) on NOACs. After adjustment, warfarin was not associated with significantly different risk of cognitive impairment compared with no OAC (aHR 1.03 [95% CI 0.92-1.15]) while NOACs maintained a lower risk (aHR 0.82 [95% CI 0.74-0.90]; **Supplementary Table 2**).

**Figure 3 –.**
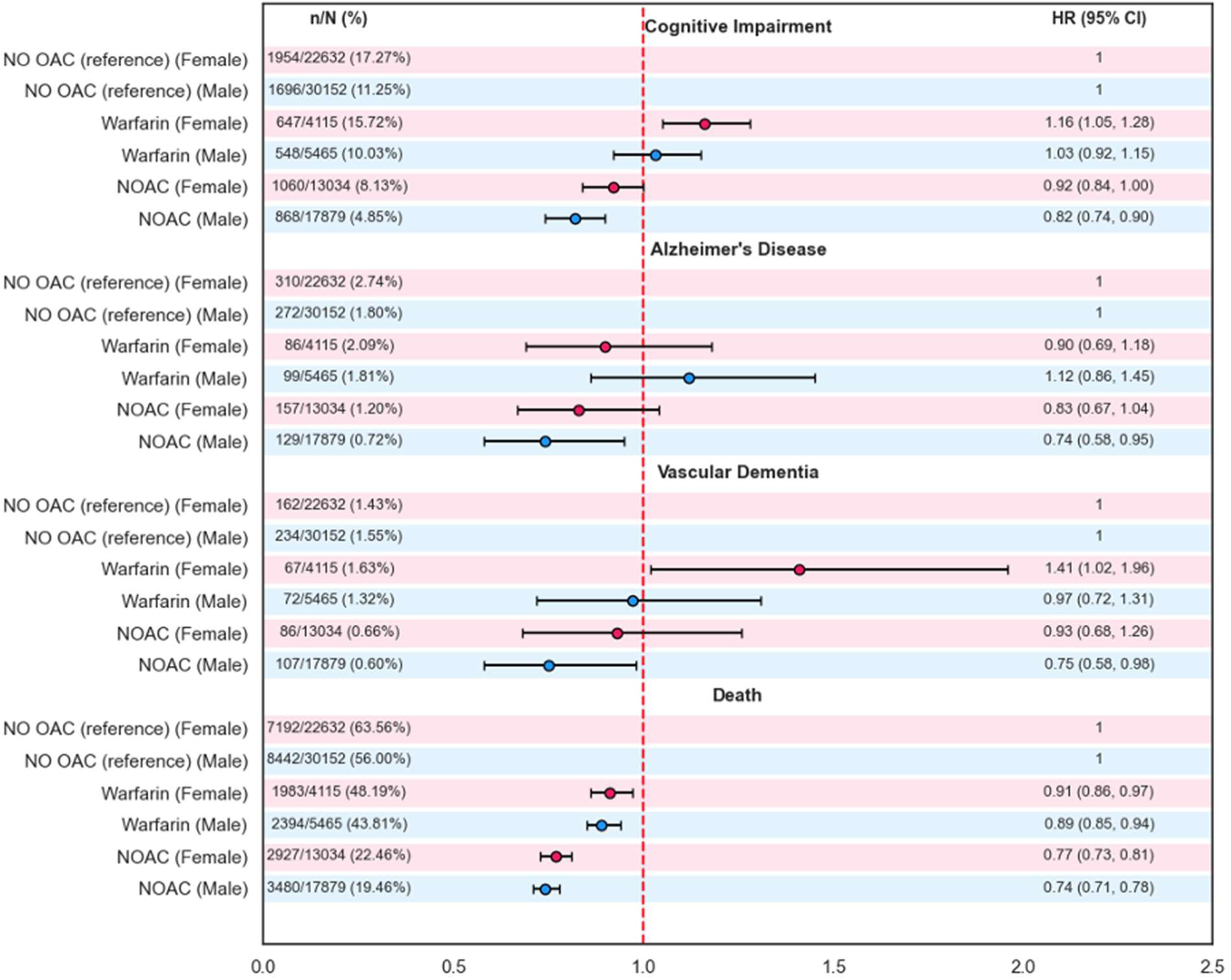
Adjusted association between oral anticoagulant (OAC) and outcomes by sex. Compared with no OAC, non-vitamin K OAC (NOAC) use was associated with lower risk of cognitive impairment of both sexes, and of Alzheimer’s disease and vascular dementia in males only. Warfarin was associated with higher risk of cognitive impairment and vascular dementia in females, but not males.

Similar patterns were observed for dementia subtypes. For Alzheimer’s disease, NOAC therapy was associated with lower risk among males (aHR 0.74 [95% CI 0.58-0.95]), whereas the association did not reach statistical significance among females (aHR 0.83 [95% CI 0.67-1.04]). Warfarin use was not significantly associated with Alzheimer’s disease risk modification in either sex.

For vascular dementia, NOAC therapy was associated with lower risk among males (aHR 0.75, 95% CI 0.58 to 0.98) but not among females (aHR 0.93, 95% CI 0.68 to 1.26). Warfarin use was associated with a higher risk of vascular dementia among females (adjusted HR 1.41, 95% CI 1.02 to 1.96) but not among males.

OAC therapy was associated with reduced all-cause mortality in both sexes. Compared with no anticoagulation, warfarin therapy was associated with lower mortality among females (aHR 0.91[95% CI 0.86-0.97]) and males (aHR 0.89 [95% CI 0.85-0.94]). NOAC therapy was associated with greater reductions in all-cause mortality risk, with aHR of 0.77 (95% CI 0.73-0.81) among females and 0.74 (95% CI 0.71-0.78) among males.

## Discussion

In this large population-based cohort of patients with incident NVAF, females had higher rates of cognitive impairment, Alzheimer’s disease, and all-cause mortality than males, with similar rates of vascular dementia. After adjustment, female sex remained associated with a modest increase in cognitive impairment, but not Alzheimer’s disease, and with lower risks of vascular dementia and all-cause mortality. Cognitive impairment was associated with lower risk among patients receiving NOAC therapy in both sexes, whereas warfarin was not associated with risk reduction in either males or females. Alzheimer’s disease and vascular dementia were associated with lower risk among males receiving NOAC therapy only, with no significant association observed in females; warfarin was not associated with reduced risk in either sex, and increased vascular dementia risk in females only. All-cause mortality was lower among patients treated with NOACs in both sexes, whereas warfarin was associated with only a modest reduction in mortality relative to no OAC, indicating that sex differences in AF-related cognitive outcomes vary by outcome and are influenced by baseline clinical characteristics and OAC type.

Our findings align with prior studies showing that females with AF may experience greater cognitive decline than males. Wood et al. reported higher risk of progression from normal cognition to mild cognitive impairment and vascular dementia in females in a retrospective registry, evaluating longitudinal neuropsychological testing over four years.^15^ Golive et al found that, despite lower AF incidence, females had higher long-term dementia rates in the Intermountain registry of patients who underwent coronary angiography and were followed up over at least five years, while Clúa-Espuny et al showed higher cognitive impairment prevalence in females despite greater cardiometabolic burden in males in their observational, retrospective analysis of patients in a primary care setting.^3, 19^ Our results extend these observations by demonstrating that females continue to have higher crude rates of post-diagnosis cognitive decline despite similar OAC initiation.

The attenuation of the association with Alzheimer’s disease after adjustment suggests that much of the excess risk may be explained by age and frailty rather than sex itself. Females were older, frailer, and had higher hypertension burden and CHA₂DS₂-VASc scores, whereas males had more diabetes and atherosclerotic cardiovascular disease, indicating distinct baseline risk profiles. In contrast, the persistence of a modest association with overall cognitive impairment suggests greater susceptibility among females to early or nonspecific cognitive decline not fully captured by dementia subtypes.

Several mechanisms may explain these findings. AF may contribute to cognitive decline through stroke, silent cerebral ischemia, cerebral hypoperfusion, inflammation, and cumulative hemodynamic instability.^1, 20–22^ Females in our cohort were older and frailer, with greater hypertension burden, factors that may increase vulnerability to small vessel disease and reduce cognitive reserve at diagnosis. Prior studies also suggest that females may experience delays in AF recognition and referral, potentially leading to longer untreated exposure before diagnosis.^14^ Although not directly assessed, these factors may contribute to the higher rates of cognitive impairment observed in females despite adjustment.

Compared with no OAC therapy, NOAC use was associated with lower risk of cognitive impairment in both sexes, and of Alzheimer’s disease and vascular dementia in males. In contrast, warfarin was associated with higher risk of cognitive impairment and vascular dementia in females, with no significant differences in males. These findings are consistent with prior systematic reviews showing that anticoagulation may reduce dementia risk in AF and that effective stroke prevention is central to brain protection in this population.^9, 23^ Although one sub-study of ALONE-AF demonstrated no difference in cognitive impairment outcomes between those on and off OAC after AF ablation, and no significant sex differences, this was underpowered to detect differences and follow up was short at two years.^24^ Several factors may explain the more favourable associations observed with NOACs in our study. NOACs provide more predictable anticoagulation and avoid the variability seen with warfarin, potentially reducing both overt and subclinical thromboembolic injury.^9, 23, 25^ In an Alberta emergency department cohort, Islam et al showed that over half of patients on warfarin had suboptimal anticoagulation control (time in therapeutic range <65%), while NOAC adherence, particularly among females, was higher.^16^ Similarly, Yogasundaram et al reported consistently high NOAC adherence across care settings, whereas warfarin control remained suboptimal and was associated with worse outcomes.^26^ Our findings may therefore reflect not only pharmacologic differences, but also variation in real-world treatment quality not captured by OAC initiation alone.

Our study is novel by providing additional information on sex differences in cognitive impairment by stratifying the effects of OAC into cognitive outcomes in each sex. Although OAC initiation within 120 days was similar between sexes, prior Alberta data show that females presenting with incident AF were less likely to receive OAC at discharge and had higher one-year stroke risk. More broadly, NOAC uptake and adherence varied by care setting, with higher rates in outpatient compared with inpatient or emergency settings.^16, 26^ The absence of a difference in our study should not be interpreted as treatment equity across the AF care continuum, as differences in diagnosis setting, timing, and adherence may still contribute to sex-specific cognitive outcomes.

### Limitations

This was an observational study using administrative data, limiting causal inference and despite multivariable adjustment, residual confounding remains possible. We lacked data on baseline cognitive function, AF burden, neuroimaging, and detailed rhythm-control therapies, all of which may influence cognitive decline. Outcomes were identified using diagnostic codes rather than standardized neuropsychological assessment, which may have led to under-ascertainment of mild cognitive impairment and potential detection bias, particularly in older or frailer patients. Dementia subtype classification is especially prone to misclassification in administrative data. Finally, anticoagulant analyses were based on treatment initiation and did not account for adherence, quality of anticoagulation control, or switching over time. Females receiving warfarin may have differed from those receiving NOACs in ways not fully captured, and residual confounding by indication is likely, especially if warfarin was used for more complex or frail patients, of whom more were females.

## Conclusion

In this large population-based study, we found females have a higher risk of cognitive impairment, but a lower risk of vascular dementia and all-cause mortality compared to males. For both sexes, NOAC use was associated with a lower risk of cognitive impairment compared to no OAC and a lower risk of Alzheimer’s disease and vascular dementia in males only.

## Data Availability

All data used for formulation of this manuscript is available upon request

## Acknowledgements

This study is based in part on data provided by PPHS (Primary & Preventative Health Services) and Alberta Health Services. We thank the Customer Relationship Management and Data Access Unit at PPHS for creating the linked database. The interpretation and conclusions are those of the researchers and do not represent the views of the Government of Alberta. Neither the Government of Alberta nor PPHS express any opinion in relation to this study.

